# Factors Associated with Physical Function among People with Systemic Sclerosis: A Scleroderma Patient-centered Intervention Network (SPIN) Cohort Cross-sectional Study

**DOI:** 10.1101/2023.08.23.23294495

**Authors:** Tiffany Dal Santo, Danielle B. Rice, Marie-Eve Carrier, Gabrielle Virgili-Gervais, Brooke Levis, Linda Kwakkenbos, Susan J. Bartlett, Amy Gietzen, Karen Gottesman, Geneviève Guillot, Marie Hudson, Laura K. Hummers, Vanessa L. Malcarne, Maureen D. Mayes, Luc Mouthon, Michelle Richard, Maureen Sauvé, Robyn K. Wojeck, Marie-Claude Geoffroy, Andrea Benedetti, Brett D. Thombs, the SPIN investigators

## Abstract

**Objectives:** To compare physical function in systemic sclerosis (SSc, scleroderma) to general population normative data and identify associated factors.

**Methods:** Scleroderma Patient-centered Intervention Network Cohort participants completed the Physical Function domain of the Patient-Reported Outcomes Measurement Information System Version 2 upon enrollment. Multivariable linear regression was used to assess associations of sociodemographic, lifestyle, and disease-related variables.

**Results:** Among 2,385 participants, mean physical function T-score (43.7, SD = 8.9) was approximately 2/3 of a standard deviation (SD) below the US general population (mean = 50, SD = 10). Factors associated in multivariable analysis included older age (-0.74 points per SD years, 95% CI -0.78 to -1.08), female sex (-1.35, -2.37 to -0.34), fewer years of education (-0.41 points per SD in years, -0.75 to -0.07), being single, divorced, or widowed (-0.76, -1.48 to -0.03), smoking (-3.14, -4.42 to -1.85), alcohol consumption (0.79 points per SD drinks per week, 0.45 to 1.14), BMI (-1.41 points per SD, -1.75 to -1.07), diffuse subtype (-1.43, -2.23 to -0.62), gastrointestinal involvement (-2.58, -3.53 to -1.62), digital ulcers (-1.96, -2.94 to -0.98), moderate (-1.94, -2.94 to -0.93) and severe (-1.76, -3.24 to -0.28) small joint contractures, moderate (-2.10, -3.44 to -0.76) and severe (-2.54, -4.64 to -0.44) large joint contractures, interstitial lung disease (-1.52, -2.27 to -0.77); pulmonary arterial hypertension (-3.72, -4.91 to -2.52); rheumatoid arthritis (-2.10, -3.64 to -0.56) and idiopathic inflammatory myositis (-2.10, -3.63 to -0.56).

**Conclusion:** Physical function is impaired for many individuals with SSc and associated with multiple disease factors.

**KEY MESSAGES:**

- Individuals with systemic sclerosis (SSc) face many challenges that can impact their physical function
- Levels of physical function in individuals with SSc are impaired compared to the general population
- Multiple disease factors are significantly associated with worse physical function in SSc

---

Systemic sclerosis (SSc, scleroderma) is a complex, rare, chronic autoimmune disease involving microvascular damage that is characterized by fibrosis of the skin and other organs, including the lungs, gastrointestinal tract, kidneys, and heart.^1^ Challenges that negatively impact health-related quality of life include disability and diminished physical function, respiratory difficulty, gastrointestinal symptoms, fatigue, pain, sleep disruptions, body image distress, uncertainty and fear of disease progression, and symptoms of depression and anxiety.^1^

Physical function in SSc appears to be substantially impaired. A 2009 systematic review (12 studies; 1,127 participants) estimated that the physical component summary (PCS) score of the Short Form Survey-36 among people with SSc was 38 (95% confidence interval (CI) 35 to 42), which is 1.2 standard deviations (SDs) below the US general population mean.^2^ A 2018 systematic review of studies that compared people with SSc and healthy controls (7 studies; 795 patients and 1,154 healthy controls) reported a pooled mean PCS score of 41 (95% CI 31 to 53) in SSc, which was 15 points (95% CI 11 to 19 points) lower than control participants.^3^

Only one study with at least 200 participants has examined factors potentially associated with physical function in SSc. That study evaluated 578 participants from the Canadian Scleroderma Research Group Registry and found that age, modified Rodnan Skin Score (mRSS), tender joints, gastrointestinal symptoms, breathing problems, pruritus, and Raynaud’s phenomenon were significantly related to SF-36 PCS scores. However, many key disease-related factors were subjectively reported by patients (e.g., number of gastrointestinal symptoms, severity of breathing problems, severity of Raynaud’s phenomenon), which may have magnified associations with self-reported physical function compared to objectively assessed disease status indicators.^4^

A better understanding of physical function in SSc and associated disease manifestations would support research on approaches to disease management to improve quality of life. Our objective was to (1) compare physical function levels in a large multinational SSc cohort to general population normative data and 2) identify sociodemographic, lifestyle and SSc disease factors associated with physical function.

## METHODS

This was a cross-sectional study that evaluated baseline data from the Scleroderma Patient-centered Intervention Network (SPIN) Cohort.^5–7^ It was reported based on guidance in the Strengthening the Reporting of Observational Studies in Epidemiology Statement.^8^ Methods from studies that use data from the SPIN Cohort are similar. Thus, we followed reporting guidance from the Text Recycling Research Project.^9^

### Participants and Procedures

The SPIN Cohort is a convenience sample of participants from 7 countries: Australia, Canada, France, Mexico, the United Kingdom, the United States, and Spain.^5–7^ Eligible participants are recruited by the attending physician or a nurse coordinator during regular physician visits. Participants included in the SPIN Cohort must be ≥18 years of age; fluent in English, French or Spanish; and classified as having SSc based on the 2013 American College of Rheumatology/European League Against Rheumatism classification criteria for SSc^10^ as verified by a SPIN site physician. After obtaining written informed consent from eligible participants, onsite staff submit an online medical data form and participants receive an automated email with instructions on how to activate their online SPIN account and complete their baseline measures. SPIN Cohort participants complete subsequent online assessments every 3 months. The SPIN Cohort study was approved by the Research Ethics Committee of the Centre intégré universitaire de santé et de services sociaux du Centre-Ouest-de-l’Île-de-Montréal (#MP-05-2013-150) and by the ethics committees of all recruiting sites. Participant recruitment is ongoing. This study used baseline assessment data from participants enrolled in the SPIN Cohort from April 2014, the date of inception, until March 2023. SPIN Cohort participants were included in this study if they completed all Patient Reported Outcomes Information System (PROMIS-29) version 2.0 domains at their baseline assessment.

### Measures

SPIN Cohort participants provided sociodemographic (race or ethnicity, education level, marital status) and lifestyle (e.g., smoking status, alcohol consumption) information and completed patient-reported outcome measures. Physicians reported participants’ age; sex; height; weight; years since initial onset of non-Raynaud phenomenon symptoms; SSc subtype (limited, diffuse, sine); mRSS; presence of gastrointestinal symptoms (upper; lower; or no gastrointestinal involvement); presence of digital ulcers anywhere on the fingers; presence of tendon friction rubs (currently; in the past; never); presence of small or large joint contractures (none; mild [≤ 25% range of motion limitation]; moderate to severe [> 25%]); presence of pulmonary arterial hypertension; presence of interstitial lung disease; existing history of SSc renal crisis; presence of current or past overlap syndromes (systemic lupus erythematosus, rheumatoid arthritis, Sjogren’s syndrome, autoimmune thyroid disease, idiopathic inflammatory myositis, and primary biliary cirrhosis); and presence of SSc-related antibodies (antinuclear antibody, anti-centromere, anti-topoisomerase I and anti-RNA polymerase III).

#### Physical Function

Physical function was evaluated using the 4a Short Form of the PROMIS-29 v2.0 Physical Function domain, which assesses patient-reported health status over the past 7 days.^11^ Item scores are summed to give a Physical Function domain score that is converted into a T-score normalized to the United States general population (mean = 50, SD = 10).^12^ A normal level of physical functioning is represented by a T-score over 45.0, mild impairment by a T-score between 40.0 to 45.0, moderate impairment between 30.0 to 39.9 and severe impairment in physical functioning by a T-score less than 30.0.^13^ The PROMIS-29v2.0 has been validated within the SPIN Cohort, with Cronbach’s α ranging from 0.86 to 0.96 for all PROMIS-29v2.0 domains and good convergent validity.^14^

#### Pruritus

Pruritus severity was evaluated with a single item: “In the past week, how severe was your itch?”, with patients using a 11-point numeric rating scale (0 = not severe at all to 11 = unbearable). Similar numerical rating scales have been shown to be valid for assessing pruritus severity.^15^

#### Pain

Pain intensity in the last week was assessed with the PROMIS-29v2.0 using a single-item: “In the past seven days, how would you rate your pain on average?”.^16,17^ This item is rated on a 10-point numerical rating scale (0 = no pain to 10 = worst imaginable pain). Single- and multi-level item measurements of pain intensity have been shown to perform equivalently in individuals with SSc.^18^ Pain interference in the last week was assessed with the PROMIS-29v2.0 using 4 items, each rated on a 5-point Likert scale (1 = “Not at all” and 5 = “Very much”).

### Statistical Analysis

We computed descriptive statistics for all variables for the entire sample and separately for those with diffuse and limited SSc (including sine) and by sex. Unadjusted outcomes were generated with simple linear regressions used to assess bivariate associations of sociodemographic, lifestyle, and disease-related variables with physical function. Adjusted outcomes were generated with multivariable linear regression used to assess the independent association of each variable with physical function. We identified items to be included in the model *a priori* based on previous studies of factors associated with physical function and other patient-reported outcomes in SSc^4,19–22^ and on the experience of research team members who either have or provide health care for individuals with SSc. We did not include psychosocial or functional variables that are outcomes of SSc (depression symptoms, anxiety symptoms, pain, fatigue, self-efficacy) as predictors in the main model as they are likely to have bidirectional causal associations with physical function. We did this to avoid reverse causality where outcome variables may be causally associated to predictor variables, which can lead to (1) biased model coefficients, potentially masking important associations between disease variables and physical function; (2) spuriously inflated goodness-of-fit estimates (R^2^); and (3) inability to determine the relative causal influence between the variables for which reverse causation is likely.^23^

Variables included in the main analysis were age (years standardized); male sex (reference = female); years of education (years standardized); single, divorced/separated, or widowed (reference = married or living as married); non-White (reference = White); Canada, United Kingdom, France, other (Australia, Mexico, Spain) (reference = United States); smoker (reference = non-smoker); alcohol consumption (drinks per week standardized); body mass index (BMI) (standardized); years since first non-Raynaud’s symptoms (years standardized); diffuse subtype (reference = limited or sine); gastrointestinal involvement (reference = no); digital ulcers (reference = no); current or past tendon friction rubs (reference = never); moderate or severe small joint contractures (reference = none or mild); moderate or severe large joint contractures (reference = none or mild); history of SSc renal crisis (reference = no); interstitial lung disease (reference = no); pulmonary arterial hypertension (reference = no); systemic lupus erythematosus (reference = no); rheumatoid arthritis (reference = no); Sjogren’s syndrome (reference = no); autoimmune thyroid disease (reference = no); idiopathic inflammatory myositis (reference = no); primary biliary cirrhosis (reference = no). See Supplementary Table S1 for variable specifications.

We accounted for missing data by using multiple imputation via chained equations, using the mice package in R.^24^ We generated 20 imputed datasets, using 15 cycles per dataset. Variables included in the mice procedure included: all variables in the main regression model, all variables considered in sensitivity analyses, and anxiety, depression, pain intensity and interference, fatigue, sleep, and satisfaction with social roles and activities function domain scores on the PROMIS-29v2.0.

We conducted 4 multivariable sensitivity analyses. We (1) conducted a complete case analysis of the main model; (2) added pruritus and pain to the main model since the direction of the association of pain and pruritus with physical function was hypothesized to be predominantly from pain and pruritis towards physical function; (3) replaced disease subtype with continuous mRSS; and (4) added SSc-related antibodies (antinuclear antibodies (reference = negative); anti-centromere (reference = negative); anti-topoisomerase I (reference = negative); and anti-RNA polymerase III (reference = negative)) to the main model. See Supplementary Table S1.

We standardized continuous predictor variables after imputation and prior to entering them in the models. We reported unstandardized regression coefficients with 95% CIs and total explained variance for each model (adjusted *R*^*2*^). All regression analyses were conducted in R (R version 3.6.3, RStudio Version 1.2.5042).

### Patient involvement

Patient members of the SPIN Steering Committee play a role in developing SPIN research priorities, including identifying the need for the present study. Five patient members of the Steering Committee reviewed and provided comments on the study protocol and manuscript and are co-authors.

## RESULTS

Our sample consisted of 2,385 participants from 53 sites with baseline PROMIS-29v2.0 Physical Function domain scores. Participants were predominantly female (N= 2,079; 87%) and White (N= 1,970; 83%). Mean (SD) age was 54.9 (12.6) years, mean (SD) education was 15.0 (3.7) years, and mean (SD) BMI was 25.3 (5.6). Most participants were from the United States (N=813; 34%), France (N=713; 30%), or Canada (N=515; 22%). Mean (SD) time in years since onset of first non-Raynaud’s symptoms was 10.9 (8.8), and 904 (38%) participants had diffuse SSc. Table 1 shows participant sociodemographic and disease characteristics, including the number with data for each variable, for the full sample and by disease subtype. See Supplementary Table S2 for participant characteristics by sex.

**Table 1.** Sample sociodemographic and disease characteristics.

|  | Full sample<br>(N = 2,385) |  | Limited SSc<sup>a</sup> (N = 1,456) |  | Diffuse SSc (N = 904) |  |
| --- | --- | --- | --- | --- | --- | --- |
|  | N<sup>b</sup> | Mean (SD) or N (%) | N | Mean (SD) or N (%) | N | Mean (SD) or N (%) |
| Age (years) | 2,381 | 54.9 (12.6) | 1,452 | 56.6 (12.4) | 904 | 52.1 (12.4) |
| Sex | 2,385 |  | 1,456 |  | 904 |  |
| Female |  | 2,079 (87%) |  | 1,299 (89%) |  | 759 (84%) |
| Male |  | 306 (13%) |  | 157 (11%) |  | 145 (16%) |
| Education (years) | 2,377 | 15.0 (3.7) | 1,450 | 14.9 (3.8) | 902 | 15.1 (3.6) |
| Marital status | 2,381 |  | 1,453 |  | 903 |  |
| Married or living as married |  | 1,661 (70%) |  | 1,035 (71%) |  | 611 (68%) |
| Single, divorced/separated, widowed |  | 720 (30%) |  | 418 (29%) |  | 292 (32%) |
| Race or ethnicity | 2,379 |  | 1,453 |  | 901 |  |
| White |  | 1,970 (83%) |  | 1,268 (87%) |  | 684 (76%) |
| Non-white |  | 409 (17%) |  | 185 (13%) |  | 217 (24%) |
| Country | 2,383 |  | 1,455 |  | 903 |  |
| United States |  | 813 (34%) |  | 436 (30%) |  | 370 (41%) |
| France |  | 713 (30%) |  | 470 (32%) |  | 241 (27%) |
| Canada |  | 515 (22%) |  | 332 (23%) |  | 173 (19%) |
| United Kingdom |  | 241 (10%) |  | 143 (10%) |  | 94 (10%) |
| Australia, Mexico, Spain |  | 101 (4%) |  | 74 (5%) |  | 25 (3%) |
| Smoking status | 2,382 |  | 1,454 |  | 903 |  |
| Smoker |  | 177 (7%) |  | 118 (8%) |  | 55 (6%) |
| Non-smoker |  | 2,205 (93%) |  | 1,336 (92%) |  | 848 (94%) |
| Alcohol consumption (drinks per week) | 2,379 | 2.0 (4.1) | 1,453 | 2.1 (4.3) | 901 | 1.8 (3.5) |
| Body mass index | 2,385 | 25.3 (5.6) | 1,456 | 25.5 (5.6) | 904 | 24.9 (5.7) |
| Years since first non-Raynaud's symptoms | 2,190 | 10.9 (8.8) | 1,325 | 12.1 (9.3) | 843 | 8.9 (7.4) |
| Disease subtype | 2,360 |  | 1,456 |  | 904 |  |
| Diffuse |  | 904 (38%) |  | 0 (0%) |  | 904 (100%) |
| Limited or sine<sup>a</sup> |  | 1,456 (62%) |  | 1,456 (100%) |  | 0 (0%) |
| mRSS | 1,983 | 7.7 (8.0) | 1,213 | 4.2 (4.2) | 754 | 13.4 (9.4) |
| Gastrointestinal involvement | 2,353 |  | 1,442 |  | 891 |  |
| Yes |  | 2,016 (85%) |  | 1,225 (85%) |  | 779 (87%) |
| No |  | 337 (14%) |  | 217 (15%) |  | 112 (13%) |
| Digital ulcers | 2,287 |  | 1,407 |  | 859 |  |
| Yes |  | 365 (16%) |  | 132 (9%) |  | 228 (27%) |
| No |  | 1,922 (84%) |  | 1,275 (91%) |  | 631 (73%) |
| Tendon friction rubs | 2,100 |  | 1,308 |  | 775 |  |
| Current |  | 232 (11%) |  | 120 (9%) |  | 110 (14%) |
| Past |  | 221 (11%) |  | 41 (3%) |  | 178 (23%) |
| Never |  | 1,647 (78%) |  | 1,147 (88%) |  | 487 (63%) |
| Small Joint Contractures | 2,255 |  | 1,388 |  | 848 |  |
| None or mild |  | 1,663 (74%) |  | 1,176 (85%) |  | 473 (56%) |
| Moderate |  | 424 (19%) |  | 161 (12%) |  | 260 (31%) |
| Severe |  | 168 (7%) |  | 51 (4%) |  | 115 (14%) |
| Large Joint Contractures | 2,211 |  | 1,359 |  | 833 |  |
| None or mild |  | 1,937 (88%) |  | 1,263 (93%) |  | 658 (79%) |
| Moderate |  | 200 (9%) |  | 65 (5%) |  | 134 (16%) |
| Severe |  | 74 (3%) |  | 31 (2%) |  | 41 (5%) |
| History of SSc renal crisis | 2,351 |  | 1,442 |  | 890 |  |
| Yes |  | 101 (4%) |  | 24 (2%) |  | 77 (9%) |
| No |  | 2,250 (96%) |  | 1,418 (98%) |  | 813 (91%) |
| Interstitial lung disease | 2,335 |  | 1,431 |  | 883 |  |
| Yes |  | 827 (35%) |  | 390 (27%) |  | 432 (49%) |
| No |  | 1,508 (65%) |  | 1,041 (73%) |  | 451 (51%) |
| Pulmonary arterial hypertension | 2,271 |  | 1,398 |  | 852 |  |
| Yes |  | 207 (9%) |  | 131 (9%) |  | 74 (9%) |
| No |  | 2,064 (91%) |  | 1,267 (91%) |  | 778 (91%) |
| Pruritus | 2,154 | 1.8 (2.6) | 1,300 | 1.6 (2.5) | 831 | 2.1 (2.8) |
| Pain intensity | 2,385 | 3.6 (2.6) | 1,456 | 3.5 (2.6) | 904 | 3.8 (2.6) |
| Pain interference | 2,384 | 55.5 (9.7) | 1,455 | 54.8 (9.6) | 904 | 56.6 (9.7) |
| Systemic lupus erythematosus | 2,323 |  | 1,428 |  | 876 |  |
| Yes |  | 65 (3%) |  | 44 (3%) |  | 20 (2%) |
| No |  | 2,258 (97%) |  | 1,384 (97%) |  | 856 (98%) |
| Rheumatoid arthritis | 2,322 |  | 1,426 |  | 877 |  |
| Yes |  | 125 (5%) |  | 64 (4%) |  | 59 (7%) |
| No |  | 2,197 (95%) |  | 1,362 (96%) |  | 818 (93%) |
| Sjogren's syndrome | 2,285 |  | 1,403 |  | 863 |  |
| Yes |  | 176 (8%) |  | 124 (9%) |  | 52 (6%) |
| No |  | 2,109 (92%) |  | 1,279 (91%) |  | 811 (94%) |
| Autoimmune thyroid disease | 2,277 |  | 1,397 |  | 861 |  |
| Yes |  | 143 (6%) |  | 99 (7%) |  | 44 (5%) |
| No |  | 2,134 (94%) |  | 1,298 (93%) |  | 817 (95%) |
| Idiopathic inflammatory myositis | 2,322 |  | 1,430 |  | 872 |  |
| Yes |  | 121 (5%) |  | 60 (4%) |  | 59 (7%) |
| No |  | 2,201 (95%) |  | 1,370 (96%) |  | 813 (93%) |
| Primary biliary cirrhosis | 2,301 |  | 1,413 |  | 869 |  |
| Yes |  | 44 (2%) |  | 38 (3%) |  | 5 (1%) |
| No |  | 2,257 (98%) |  | 1,375 (97%) |  | 864 (99%) |
| Antinuclear antibodies | 2,194 |  | 1,360 |  | 818 |  |
| Positive |  | 2,069 (94%) |  | 1,296 (95%) |  | 757 (93%) |
| Negative |  | 125 (6%) |  | 64 (5%) |  | 61 (7%) |
| Anti-centromere | 1,861 |  | 1,171 |  | 680 |  |
| Positive |  | 665 (36%) |  | 609 (52%) |  | 54 (8%) |
| Negative |  | 1,196 (64%) |  | 562 (48%) |  | 626 (92%) |
| Anti-topoisomerase I [Scl70] | 2,077 |  | 1,267 |  | 799 |  |
| Positive |  | 555 (27%) |  | 243 (19%) |  | 311 (39%) |
| Negative |  | 1,522 (73%) |  | 1,024 (81%) |  | 488 (61%) |
| Anti-RNA polymerase III | 1,353 |  | 808 |  | 539 |  |
| Positive |  | 245 (18%) |  | 49 (6%) |  | 195 (36%) |
| Negative |  | 1,108 (82%) |  | 759 (94%) |  | 344 (64%) |

<sup>a</sup>Includes 73 participants with sine SSc; <sup>b</sup>N for some variables < 2,385 due to missing data.

As shown in Table 2, the mean (SD) physical function score in the full sample was 43.7 (8.9), which is considerably lower than the United States general population mean (SD) of 50 (10). Among all participants, 1,005 (42%) had physical function scores within normal limits (T-score > 45); 508 (21%) reported mild impairment (T-score 40 to 45), 787 (33%) moderate impairment (T-score 30 to 39.9); and 85 (4%) severe impairment (T-score < 30). By country, mean (SD) scores ranged from 41.8 (9.9) among 241 participants from the UK to 45.5 (8.4) in 101 participants from Australia, Mexico, or Spain. Participants with diffuse SSc reported somewhat lower mean (SD) physical function scores (42.0 [8.4]) than those with limited or sine SSc (44.8 [8.9]). Scores for females and males were similar.

**Table 2.** Physical function by country, disease subtype, and sex.

|  | T-score mean | Within normal limits<br>(T-score > 45) | Mild<br>(T-score 40 to 45) | Moderate<br>(T-score 30 to 39.9) | Severe<br>(T-score < 30) |
| --- | --- | --- | --- | --- | --- |
| Full sample (N = 2,385) | 43.7 (8.9) | 1,005 (42%) | 508 (21%) | 787 (33%) | 85 (4%) |
| Country |  |  |  |  |  |
| USA (N = 813) | 43.6 (8.6) | 331 (41%) | 186 (23%) | 274 (34%) | 22 (3%) |
| France (N = 713) | 44.2 (8.6) | 319 (45%) | 153 (22%) | 223 (31%) | 18 (3%) |
| Canada (N = 515) | 43.7 (9.1) | 220 (43%) | 105 (20%) | 169 (33%) | 21 (4%) |
| UK (N = 241) | 41.8 (9.9) | 88 (37%) | 36 (15%) | 94 (39%) | 23 (10%) |
| Other<sup>a</sup> (N = 101) | 45.5 (8.4) | 47 (47%) | 27 (27%) | 26 (26%) | 1 (1%) |
| SSc Subtype |  |  |  |  |  |
| Limited or sine<sup>b</sup> (N = 1,456) | 44.8 (8.9) | 681 (47%) | 302 (21%) | 429 (30%) | 44 (3%) |
| Diffuse (N = 904) | 42.0 (8.4) | 317 (35%) | 202 (22%) | 348 (39%) | 37 (4%) |
| Sex |  |  |  |  |  |
| Female (N = 2,079) | 43.6 (8.9) | 878 (42%) | 436 (21%) | 688 (33%) | 77 (4%) |
| Male (N = 306) | 44.4 (8.9) | 127 (42%) | 72 (24%) | 99 (32%) | 8 (0%) |

<sup>a</sup>Includes 40 participants in Australia, 21 in Mexico, and 40 in Spain; <sup>b</sup> Includes 73 participants with sine SSc

In the main multivariable analysis (Table 3), among sociodemographic variables, older age (-0.74 points per SD in years, 95% CI -0.78 to -1.08); female sex (-1.35 points, 95% CI -2.37 to -0.34); fewer years of education (-0.41 points per SD in years, 95% CI -0.75 to -0.07); and being single, divorced or separated, or widowed (-0.76 points, 95% CI -1.48 to -0.03) were associated with lower physical function. Among lifestyle variables, there were significant associations with smoking (-3.14 points, 95% CI -4.42 to -1.85), alcohol consumption (0.79 points per SD in drinks per week, 95% CI 0.45 to 1.14), and BMI (-1.41 points per SD in BMI, 95% CI -1.75 to -1.07). Among disease variables, there were significant associations with diffuse subtype (-1.43 points, 95% CI -2.23 to -0.62), gastrointestinal involvement (-2.58 points, 95% CI -3.53 to -1.62), digital ulcers (-1.96 points, 95% CI -2.94 to -0.98), moderate (-1.94 points, 95% CI -2.94 to -0.93) and severe (-1.76 points, 95% CI -3.24 to -0.28) small joint contractures, moderate (-2.10 points, 95% CI -3.44 to -0.76) and severe (-2.54 points, 95% CI -4.64 to -0.44) large joint contractures, interstitial lung disease (-1.52 points, 95% CI -2.27 to -0.77), and pulmonary arterial hypertension (-3.72 points, 95% CI -4.91 to -2.52). Among overlap syndromes, rheumatoid arthritis (-2.10 points, 95% CI -3.64 to -0.56) and idiopathic inflammatory myositis (-2.10 points, 95% CI -3.63 to -0.56) were significantly associated. Variables not significantly associated were race or ethnicity, country, years since first non-Raynaud’s syndrome, presence of current or past tendon friction rubs, history of SSc renal crisis, systemic lupus erythematosus, Sjogren’s syndrome, autoimmune thyroid disease, and primary biliary cirrhosis. Adjusted R^2^ for the final model was 0.17.

**Table 3.**
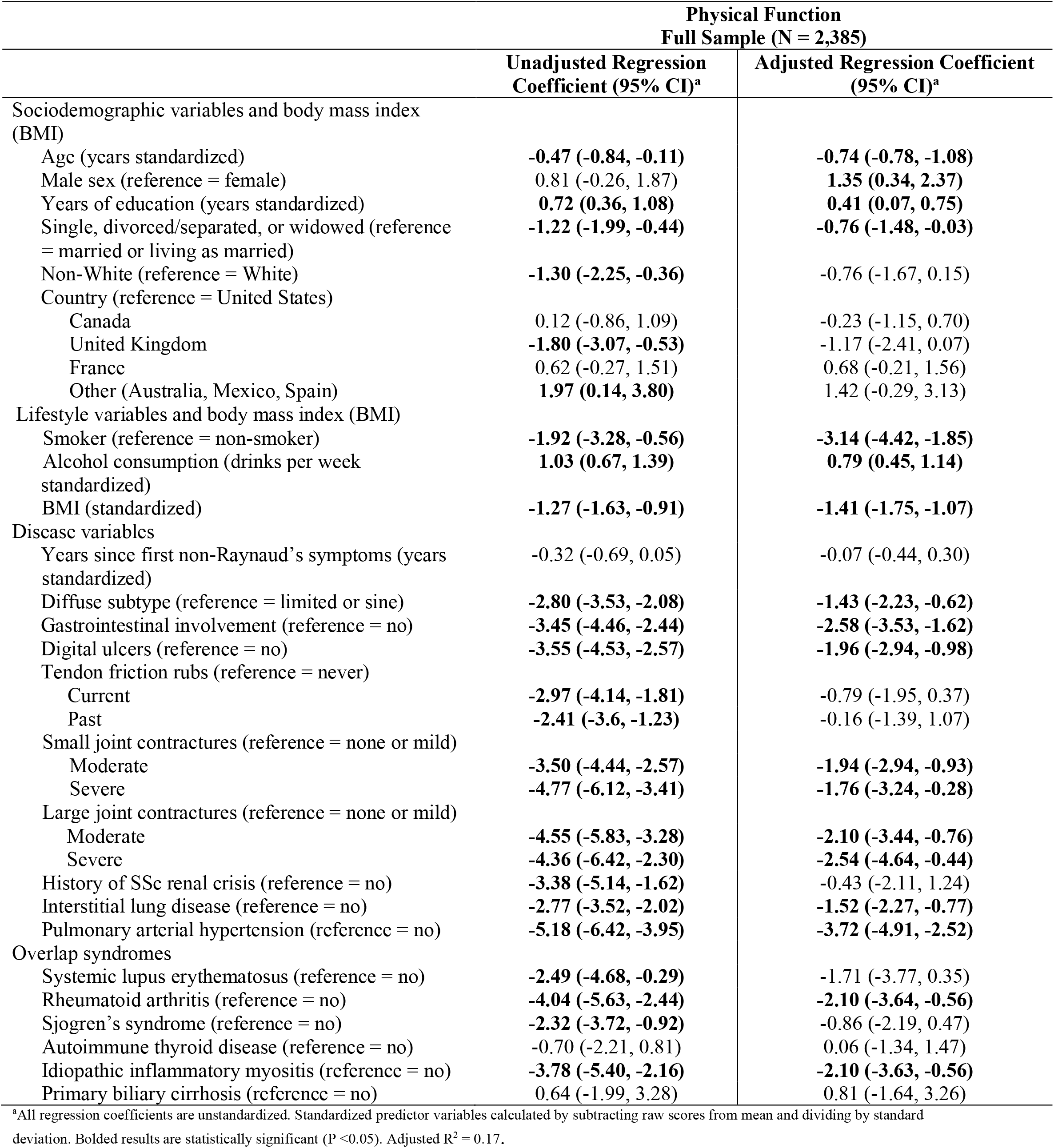
Linear regression analysis of sociodemographic and disease characteristic associations with physical function.

In sensitivity analyses, complete case analysis results, which included 1,663 participants, were similar to those of main analyses (see Supplementary Table S3). When adding pruritus and pain intensity to the model, both pruritus (-0.68 points per SD in pruritus, 95% CI -1.00 to -0.37) and pain intensity (-4.55 points per SD in pain intensity, 95% CI -4.87 to -4.24) were associated with lower physical function. In the analysis that used mRSS instead of disease subtype, mRSS was significantly associated with worse physical function (-0.99 points per SD in mRSS score, 95% CI -1.41 to -0.58). Lastly, when adding SSc-related antibodies to the model, we found Anti-topoisomerase I [Scl70] (positive) to have a significant association (0.96 points, 95% CI 0.04 to 1.89). No results from other variables changed substantively in sensitivity analyses. See Supplementary Tables S4 to S6.

## DISCUSSION

Among 2,385 participants with SSc from 7 countries, the mean T-score for physical function was 43.7, which is approximately 2/3 of a SD below the US general population (mean = 50, SD = 10). There were 58% of participants with mild (21%), moderate (33%), or severely impaired (4%) physical function. We found that disease variables associated with worse physical function included diffuse SSc subtype or mRSS, gastrointestinal involvement, digital ulcers, the presence of moderate or severe small or large joint contractures, interstitial lung disease, pulmonary arterial hypertension, and the presence of overlap syndromes including rheumatoid arthritis and idiopathic inflammatory myositis. We also found that older age; female sex; fewer years of education; being single, divorced/separated, or widowed; smoking; fewer alcoholic drinks per week; and increased BMI were associated with worse physical function.

We assessed pain separately, in a sensitivity analysis, due to overlap that may occur in measuring pain and physical function. We found that pain likely plays an important role in an individual’s ability to complete physical tasks. More specifically, we found that pain was strongly negatively associated with physical function (-4.55 points per SD in pain intensity, 95% CI -4.87 to -4.24). This finding is consistent with results from a previous SPIN study on pain intensity and interference in SSc (N = 2,157), which found that 38% of participants reported moderate or severe pain intensity, and 35% reported moderate or severe pain interference with their ability to carry out daily activities.^25^

Our findings on physical function are generally consistent with two prior studies including the next largest study of people with SSc and a large study of people with other rheumatic diseases such as rheumatoid arthritis or systemic lupus erythematosus.^26,27^ A study of 477 Australian patients with SSc reported a mean (SD) on the PROMIS-29v1 Physical Function domain of 41.9 (8.6).^26^ The study of 4,346 participants with rheumatoid arthritis and 240 with systemic lupus erythematosus reported PROMIS-29 Physical Function domain means (SD) of 42.0 (9.1) for rheumatoid arthritis, and 43.9 (9.7) for systemic lupus erythametosus.^27^ No previous study has examined the association of a large number of physician-assessed SSc disease manifestations with physical function, as we did in the present study. The long list of disease factors that we found to be associated with lower physical function highlights the many challenges faced by people with SSc. SSc is a highly heterogeneous disease, but several key factors that are commonly experienced, including diffuse disease subtype, gastrointestinal involvement, and interstitial lung disease, were robustly associated with physical function.

The adjusted R^2^ for our main multivariable regression model was 0.17. This may appear low, but it is expected in samples comprised entirely of people with a chronic condition as all have the similar experience of living with the condition. High R^2^ values are important in predictive modelling, but much less so when models are used for testing hypotheses about possible associations of variables of interest with critical patient-important outcomes. In this case, including in the present study, having a sufficiently large sample size to generate reasonably precise parameter estimates is a more important consideration.^23^

Management strategies and interventions to address the high level of impairment in physical function in SSc are needed. A 2019 systematic review on the effect and safety of exercise therapy in patients with SSc only found 4 randomized controlled trials (RCTs), all with small samples (maximum N per trial arm = 16), and concluded that there was sparse evidence that could not be used to draw strong conclusions on effectiveness.^28^ A 2017 RCT (N = 220) compared a 1-month personalized physical therapy program involving trained physiotherapists and occupational therapist to usual care in patients with SSc and reported short-term effects on disability but minimal positive long-term outcomes.^29^

Self-management programs are commonly used to help people more effectively manage their disease in arthritis^28^and other common chronic conditions.^31^ In SSc, an RCT (N = 267) tested the effects of a self-administered internet-based self-management program in comparison to an educational booklet on improving self-efficacy for disease management but did not find the intervention to be statistically superior to control.^32^ Currently, SPIN is conducting a trial to compare the SPIN-SELF program, a self-management program that provides patients with essential knowledge and coping skills to help better manage day-to-day problems in SSc, to usual care.^33^ This program combines self-management modules with expert and patient instructions delivered online with support from peer-led groups.^33^

Strengths of our study include its large international sample with participants from 53 SPIN sites across 7 countries; the inclusion of a large number of sociodemographic, lifestyle, and physician-assessed disease-related factors in analyses; and the involvement of people with lived SSc experience in the project via leadership in SPIN and participation in the study. There are also limitations to consider. First, the SPIN Cohort is a convenience sample. However, a comparison with the European Scleroderma Trials and Research and Canadian Scleroderma Research Group cohorts indicated broad comparability of participant characteristics, which supports generalizability in SSc.^5^ Second, participants were required to answer questions via online questionnaires, which may potentially reduce generalizability of results. Third, our study was cross-sectional, which does not allow us to infer causality based on our results.

In summary, we found that physical function in patients with SSc is substantially impaired on average and that many factors likely contribute to this. SSc disease manifestations associated with lower physical function included disease subtype and skin score, gastrointestinal involvement, digital ulcers, small or large joint contractures, interstitial lung disease, pulmonary arterial hypertension, rheumatoid arthritis, idiopathic inflammatory myositis, as well as pruritus and pain intensity, and anti-topoisomerase I [Scl70]. Many of these had strong associations with physical function such as smoking, GI involvement, large joint contractures, pulmonary arterial hypertension, rheumatoid arthritis, and idiopathic inflammatory myositis. More studies are needed to better understand the role each of these factors play in physical function and to develop strategies that target specific factors to improve function. Meanwhile, health-care providers should work with patients to identify and address SSc-related factors that are associated with limitations in physical function and help them find ways to cope with the disease and its symptoms.

## Contributions

TDS, DBR, MEC, BL, LK, MCG, AB, BDT contributed to study conceptualization; GVG, BL to data curation; TDS, GVG, BL to formal analysis; TDS, MEC, LK, BDT to funding acquisition; TDS, DBR, MEC, LK, SJB, AG, KG, GG, MH, LKH, VM, MDM, LM, MR, MS, RW, BDT to investigation; TDS, DBR, MEC, BL, LK, BDT to methodology; MEC to project administration; DBR, MCG, AB, BDT to supervision; TDS to visualization; TDS, BDT to writing the original draft; and all authors reviewing and editing the final draft.

## Data Availability

De-identified individual participant data with a data dictionary and analysis codes that were used to generate the results reported in this article will be made available upon request to the corresponding author and presentation of a methodologically sound proposal that is approved by the Scleroderma Patient-centered Intervention Network Data Access and Publications Committee. Data requesters will need to sign a data transfer agreement.

## REFERENCES

1. Allanore Y, Simms R, Distler O, Trojanowska M, Pope J, Denton C, et al. Systemic sclerosis. Nat Rev Dis Primers 2015;1:15002. doi:10.1038/nrdp.2015.2

2. Hudson M, Thombs BD, Steele R, Panopalis P, Newton E. Health-related quality of life in systemic sclerosis: a systematic review. Arthritis Care Res 2009;61:1112–1120. doi:10.1002/art.24676

3. Li L, Cui Y, Chen S, Zhao Q, Fu T, Ji J, et al. The impact of systemic sclerosis on health-related quality of life assessed by SF-36: a systematic review and meta-analysis. Int J Rheum Dis 2018;21:1884–1893. doi:10.1111/1756-185X.13438

4. El-Baalbaki G, Razykov I, Hudson M, Bassel M, Baron M, Thombs BD. Association of pruritus with quality of life and disability in systemic sclerosis. Arthritis Care Res 2010;62:1489–1495. doi:10.1002/acr.20257

5. Dougherty DH, Kwakkenbos L, Carrier ME, Salazar G, Assassi S, Baron M, et al. The Scleroderma Patient-Centered Intervention Network Cohort: baseline clinical features and comparison with other large scleroderma cohorts. Rheumatology 2018;57:1623–1631. doi:10.1093/rheumatology/key139

6. Kwakkenbos L, Jewett LR, Baron M, Bartlett SJ, Furst D, Gottesman K, et al. The Scleroderma Patient-centered Intervention Network (SPIN) Cohort: protocol for a cohort multiple randomised controlled trial (cmRCT) design to support trials of psychosocial and rehabilitation interventions in a rare disease context. BMJ Open 2013;3. doi:10.1136/bmjopen-2013-003563

7. The Scleroderma Patient-centered Intervention Network [Internet]. [Cited 2023, August 22]. Available from: https://www.spinsclero.com/en/about

8. Vandenbroucke JP, von Elm E, Altman DG, Gotzsche PC, Mulrow CD, Pocock SJ, et al. Strengthening the Reporting of Observational Studies in Epidemiology (STROBE): explanation and elaboration. Ann Intern Med 2007;147:W163–94. doi: 10.7326/0003-4819-147-8-200710160-00010-w1

9. Text Recycling Research Project [Internet]. Best practices for researchers. [Cited 2023, August 22]. Available from: https://textrecycling.org/resources/best-practices-for-researchers/

10. van den Hoogen F, Khanna D, Fransen J, Johnson SR, Baron M, Tyndall A, et al. 2013 classification criteria for systemic sclerosis: an American College of Rheumatology/European League Against Rheumatism collaborative initiative. Ann Rheum Dis 2013;72:1747–1755. doi:10.1136/annrheumdis-2013-204424

11. Hays RD, Spritzer KL, Schalet BD, Cella D. PROMIS(®)-29 v2.0 profile physical and mental health summary scores. Qual Life Res 2018;27:1885–1891. doi:10.1007/s11136-018-1842-3

12. HealthMeasures [Internet]. Intro to PROMIS. [Cited 2023, August 22]. Available from: https://www.healthmeasures.net/explore-measurement-systems/promis/intro-to-promis

13. HealthMeasures [Internet]. PROMIS score cut points. [Cited 2023, August 22]. Available from: https://www.healthmeasures.net/score-and-interpret/interpret-scores/promis/promis-score-cut-points

14. Kwakkenbos L, Thombs BD, Khanna D, Carrier ME, Baron M, Furst D, et al. Performance of the Patient-Reported Outcomes Measurement Information System-29 in scleroderma: a Scleroderma Patient-centered Intervention Network Cohort study. Rheumatology 2017;56:1302–1311. doi:10.1093/rheumatology/kex055

15. Phan NQ, Blome C, Fritz F, Gerss J, Reich A, Ebata T, et al. Assessment of pruritus intensity: prospective study on validity and reliability of the visual analogue scale, numerical rating scale and verbal rating scale in 471 patients with chronic pruritus. Acta Derm Venereol 2012;92:502–507. doi:10.2340/00015555-1246

16. Salaffi F, Stancati A, Silvestri CA, Ciapetti A, Grassi W. Minimal clinically important changes in chronic musculoskeletal pain intensity measured on a numerical rating scale. Eur J Pain 2004;8:283–291. doi:10.1016/j.ejpain.2003.09.004

17. Woo A, Lechner B, Fu T, Wong CS, Chiu N, Lam H, et al. Cut points for mild, moderate, and severe pain among cancer and non-cancer patients: a literature review. Ann Palliat Med 2015;4:176–183. doi:10.3978/j.issn.2224-5820.2015.09.04

18. El-Baalbaki G, Lober J, Hudson M, Baron Murray, Thombs BD. Measuring pain in systemic sclerosis: comparison of the short-form McGill Pain Questionnaire versus a single-item measure of pain. J Rheumatol 2011;38:2581–2587. doi:10.3899/jrheum.110592

19. Schieir O, Thombs BD, Hudson M, Boivin JF, Steele R, Bernatsky S, et al. Prevalence, severity, and clinical correlates of pain in patients with systemic sclerosis. Arthritis Care Res 2010;62:409–417. doi:10.1002/acr.20108

20. Thombs BD, Hudson M, Taillefer SS, Baron M. Prevalence and clinical correlates of symptoms of depression in patients with systemic sclerosis. Arthritis Care Res 2008;59:504–509. doi:10.1002/art.23524

21. Thombs BD, Hudson M, Bassel M, Taillefer SS, Baron M. Sociodemographic, disease, and symptom correlates of fatigue in systemic sclerosis: evidence from a sample of 659 Canadian Scleroderma Research Group Registry patients. Arthritis Rheum 2009;61:966–973. doi:10.1002/art.24614

22. Levis B, Kwakkenbos L, Hudson M, Baron M, Thombs BD. The association of sociodemographic and objectively-assessed disease variables with fatigue in systemic sclerosis: an analysis of 785 Canadian Scleroderma Research Group Registry patients. Clin Rheumatol 2017;36:373–379. doi:10.1007/s10067-016-3501-9

23. Allison P. Multiple Regression: A Primer. Sage Publications; 1999.

24. van Buuren S, Groothuis-Oudshoorn K. mice: Multivariate Imputation by Chained Equations in R. J Stat Softw 2011;45:1–67. doi:10.18637/jss.v045.i03

25. Lee YC, Fox RS, Kwakkenbos L, Levis B, Carrier ME, Welling J, et al. Pain levels and associated factors in the Scleroderma Patient-centered Intervention Network (SPIN) Cohort: a multicentre cross-sectional study. Lancet Rheumatol 2021;3:e844–e854. doi: 10.1016/S2665-9913(21)00318-0

26. Morrisroe K, Stevens W, Huq M, Sahhar J, Ngian GS, Zochling J, et al. Validity of the PROMIS-29 in a large Australian cohort of patients with systemic sclerosis. J Scleroderma Relat Disord 2017;2:188–195. doi: 10.5301/jsrd.5000243

27. Katz P, Pedro S, Michaud K. Performance of the Patient-Reported Outcomes Measurement Information System 29-Item Profile in rheumatoid arthritis, osteoarthritis, fibromyalgia, and systemic lupus erythematosus. Arthritis Care Res 2017;69:1312–1321. doi: 10.1002/acr.23183

28. Liem SIE, Vliet Vlieland TPM, Schoones JW, de Vries-Bouwstra JK. The effect and safety of exercise therapy in patients with systemic sclerosis: a systematic review. Rheumatol Adv Pract 2019;0:1–13. doi:10.1093/rap/rkz044

29. Rannou F, Boutron I, Mouthon L, Sanchez K, Tiffreau V, Hachulla E, et al. Personalized physical therapy versus usual care for patients with systemic sclerosis: a randomized controlled trial. Arthritis Care Res 2017;69:1050–1059. doi:10.1002/acr.23098

30. Marques A, Santos E, Nikiphorou E, Bosworth A, Carmona L. Effectiveness of self-management interventions in inflammatory arthritis: a systematic review informing the 2021 EULAR recommendations for the implementation of self-management strategies in patients with inflammatory arthritis. RMD Open 2021;7:e001647. doi:10.1136/rmdopen-2021-001647

31. Coster S, Li Y, Norman I.J. Cochrane reviews of educational and self-management interventions to guide nursing practice: a review. Int J Nurs Stud 2020;110:103698. doi:10.1016/j.ijnurstu.2020.103698

32. Khanna D, Serrano J, Berrocal VJ, Silver RM, Cuencas P, Newbill SL, et al. A randomized controlled trial to evaluate an internet-based self-management program in systemic sclerosis. Arthritis Care Res 2020;71:435–447. doi:10.1002/acr.23595

33. Kwakkenbos L, Østbø N, Carrier ME, Nielson WR, Fedoruk C, Levis B, et al. Randomized feasibility trial of the Scleroderma Patient-centered Intervention Network Self-Management (SPIN-SELF) Program. Pilot Feasibility Stud 2022;8:45. doi: 10.1186/s40814-022-00994-5

