## Supplementary file for "Factors Associated with Physical Function among People with Systemic Sclerosis: A Scleroderma Patient-centered Intervention Network (SPIN) Cohort Cross-sectional Study"

**Supplementary: Table of Contents**

**Supplementary Table S1:** Variable specifications

**Supplementary Table S2**: Sample sociodemographic and disease characteristics by sex

**Supplementary Table S3:** Linear regression analysis of sociodemographic and disease characteristic associations with physical function (complete case analysis; N = 1,663)

**Supplementary Table S4:** Linear regression analysis of sociodemographic and disease characteristic associations with physical function (sensitivity analysis; N = 2,385; adding pruritus and pain intensity)

**Supplementary Table S5:** Linear regression analysis of sociodemographic and disease characteristic associations with physical function (sensitivity analysis; N = 2,385; replacing disease subtype with continuous Modified Rodnan Skin Scores)

**Supplementary Table S6:** Linear regression analysis of sociodemographic and disease characteristic associations with physical function (sensitivity analysis; N = 2,385; adding SSc-related antibodies)

**Supplementary Table S1.** Variable specifications

| Category | Variable | Regression model | Analyses |
|---|---|---|---|
| Sociodemographic variables | | | |
| | Age, years | Continuous | All analyses |
| | Sex | Dichotomous; Reference: female (versus. male) | All analyses |
| | Education, years | Continuous | All analyses |
| | Marital status | Dichotomous; Reference: married or living as married (versus. single, divorced/separated, widowed) | All analyses |
| | Race/ethnicity | Dichotomous; Reference: white (versus. non-White) | All analyses |
| | Country | Categorical; Reference: United States (versus. Canada, United Kingdom, France, other (Australia, Mexico, Spain)) | All analyses |
| Lifestyle variables and body mass index (BMI) | | | |
| | Smoking status | Dichotomous; Reference: non-smoker (versus. smoker) | All analyses |
| | Alcohol consumption, drinks/week | Continuous | All analyses |
| | PROMIS variables: anxiety, depression, sleep, fatigue, satisfaction with social roles and activities | Continuous | Imputation analysis |
| | Body mass index, BMI score | Continuous | All analyses |
| Disease variables | | | |

| | | | |
|---|---|---|---|
| | Time since first non-Raynaud's symptoms, years | Continuous | All analyses |
| | Disease subtype | Dichotomous; Reference: limited or sine SSc (versus. diffuse SSc) | Main analysis and sensitivity analysis (adding pruritus, and adding SSc-related antibodies) |
| | Modified Rodnan skin score | Continuous | Sensitivity analysis (replacing disease subtype) |
| | Gastrointestinal involvement | Dichotomous; Reference: no GI involvement (versus any GI) | All analyses |
| | Digital ulcers | Dichotomous; Reference: none (versus. anywhere on the finger) | All analyses |
| | Tendon friction rubs | Categorical; Reference: never present (versus. currently present or present in the past) | All analyses |
| | Small joint contractures | Dichotomous; Reference: none or mild contractures (versus. moderate or severe contractures) | All analyses |
| | Large joint contractures | Dichotomous; Reference: none or mild contractures (versus. moderate or severe contractures) | All analyses |
| | History of SSc Renal Crisis | Dichotomous; Reference: no (versus. yes) | All analyses |
| | Interstitial lung disease | Dichotomous; Reference: no (versus. yes) | All analyses |
| | Pulmonary arterial hypertension | Dichotomous; Reference: no (versus. yes) | All analyses |
| | Pruritus | Continuous | Sensitivity analysis |
| | Pain intensity | Continuous | Sensitivity analysis |
| | Pain interference | Continuous | Imputation analysis |
| Presence of overlap syndromes | Systemic lupus erythematosus | Dichotomous; Reference: no (versus. yes) | All analyses |
| | Rheumatoid arthritis | Dichotomous; Reference: no (versus. yes) | All analyses |

| | | | |
|---|---|---|---|
| | Sjogren's syndrome | Dichotomous; Reference: no (versus. yes) | All analyses |
| | Autoimmune thyroid disease | Dichotomous; Reference: no (versus. yes) | All analyses |
| | Idiopathic inflammatory myositis | Dichotomous; Reference: no (versus. yes) | All analyses |
| | Primary biliary cirrhosis | Dichotomous; Reference: no (versus. yes) | All analyses |
| Antibodies | Antinuclear antibodies | Dichotomous; Reference: negative (versus. positive) | Sensitivity analysis |
| | Anti-Centromere | Dichotomous; Reference: negative (versus. positive) | Sensitivity analysis |
| | Anti-topoisomerase I [Scl70] | Dichotomous; Reference: negative (versus. positive) | Sensitivity analysis |
| | Anti-RNA polymerate III | Dichotomous; Reference: negative (versus. positive) | Sensitivity analysis |

**Supplementary Table S2.** Sample sociodemographic and disease characteristics presented by sex

| | Female (N = 2,079) | | Male (N = 306) | |
|---|---|---|---|---|
| | **N** | **Mean (SD) or N (%)** | **N** | **Mean (SD) or N (%)** |
| Age (years) | 2,075 | 54.5 (12.7) | 306 | 57.2 (11.8) |
| Sex | 2,079 | | 306 | |
| Female | | 2,079 (100%) | | 0 (0%) |
| Male | | 0 (0%) | | 306 (100%) |
| Education (years) | 2,073 | 15.0 (3.7) | 304 | 14.5 (3.9) |
| Marital status | 2,075 | | 306 | |
| Married or living as married | | 1,415 (68%) | | 246 (80%) |
| Single, divorced/separated, widowed | | 660 (32%) | | 60 (20%) |
| Race or ethnicity | 2,073 | | 306 | |
| White | | 1,712 (83%) | | 258 (84%) |
| Non-white | | 361 (17%) | | 48 (16%) |
| Country | 2,077 | | 306 | |
| United States | | 704 (34%) | | 109 (36%) |
| France | | 607 (29%) | | 106 (35%) |
| Canada | | 453 (22%) | | 62 (20%) |
| United Kingdom | | 219 (11%) | | 22 (7%) |
| Australia, Mexico, Spain | | 94 (5%) | | 7 (2%) |
| Smoking status | 2,076 | | 306 | |
| Smoker | | 155 (7%) | | 22 (7%) |
| Non-smoker | | 1,921 (93%) | | 284 (93%) |
| Alcohol consumption (drinks per week) | 2,075 | 1.8 (3.6) | 304 | 3.6 (6.1) |
| Body mass index | 2,079 | 25.2 (5.8) | 306 | 25.8 (4.2) |
| Years since first non-Raynaud’s symptoms | 1,903 | 11.1 (8.9) | 287 | 9.4 (7.9) |
| Disease subtype | 2,058 | | 302 | |
| Diffuse | | 759 (37%) | | 145 (48%) |
| Limited or sine | | 1,299 (63%) | | 157 (52%) |
| mRSS | 1,723 | 7.5 (7.9) | 260 | 9.0 (8.6) |
| Gastrointestinal involvement | 2,050 | | 303 | |
| Yes | | 1,763 (86%) | | 253 (83%) |
| No | | 287 (14%) | | 50 (17%) |
| Digital ulcers | 2,001 | | 286 | |
| Yes | | 309 (15%) | | 56 (20%) |
| No | | 1,692 (85%) | | 230 (80%) |
| Tendon friction rubs | 1,825 | | 275 | |
| Current | | 194 (11%) | | 38 (14%) |
| Past | | 180 (10%) | | 41 (15%) |
| Never | | 1,451 (80%) | | 196 (71%) |
| Small Joint Contractures | 1,962 | | 293 | |
| None or mild | | 1,461 (74%) | | 202 (69%) |
| Moderate | | 354 (18%) | | 70 (24%) |
| Severe | | 147 (7%) | | 21 (7%) |
| Large Joint Contractures | 1,926 | | 285 | |
| None or mild | | 1,689 (88%) | | 248 (87%) |
| Moderate | | 170 (9%) | | 30 (11%) |
| Severe | | 67 (3%) | | 7 (2%) |

| | | | | |
|---|---|---|---|---|
| History of SSc renal crisis | 2,048 | | 303 | |
| Yes | | 84 (4%) | | 17 (6%) |
| No | | 1,964 (96%) | | 286 (94%) |
| Interstitial lung disease | 2,034 | | 301 | |
| Yes | | 666 (33%) | | 161 (53%) |
| No | | 1,368 (67%) | | 140 (47%) |
| Pulmonary arterial hypertension | 1,982 | | 289 | |
| Yes | | 172 (9%) | | 35 (12%) |
| No | | 1,810 (91%) | | 254 (88%) |
| Pruritus | 1,885 | 1.8 (2.7) | 269 | 1.5 (2.3) |
| Pain intensity | 2,079 | 3.7 (2.6) | 306 | 3.2 (2.4) |
| Pain interference | 2,078 | 55.7 (9.7) | 306 | 54.5 (9.2) |
| Systemic lupus erythematosus | 2,027 | | 296 | |
| Yes | | 59 (3%) | | 6 (2%) |
| No | | 1,968 (97%) | | 290 (98%) |
| Rheumatoid arthritis | 2,026 | | 296 | |
| Yes | | 114 (6%) | | 11(4%) |
| No | | 1,912 (94%) | | 285 (96%) |
| Sjogren’s syndrome | 1,992 | | 293 | |
| Yes | | 163 (8%) | | 13 (4%) |
| No | | 1,829 (92%) | | 280 (96%) |
| Autoimmune thyroid disease | 1,986 | | 291 | |
| Yes | | 137 (7%) | | 6 (2%) |
| No | | 1,849 (93%) | | 285 (98%) |
| Idiopathic inflammatory myositis | 2,029 | | 293 | |
| Yes | | 101 (5%) | | 20 (7%) |
| No | | 1,928 (95%) | | 273 (93%) |
| Primary biliary cirrhosis | 2,010 | | 291 | |
| Yes | | 43 (2%) | | 1 (0%) |
| No | | 1,967 (98%) | | 290 (100%) |
| Antinuclear antibodies | 1,911 | | 283 | |
| Positive | | 1,805 (94%) | | 264 (93%) |
| Negative | | 106 (6%) | | 19 (7%) |
| Anti-centromere | 1,625 | | 236 | |
| Positive | | 617 (38%) | | 48 (20%) |
| Negative | | 1,008 (62%) | | 188 (80%) |
| Anti-topoisomerase I [Scl70] | 1,804 | | 273 | |
| Positive | | 474 (26%) | | 81 (30%) |
| Negative | | 1,330 (74%) | | 192 (70%) |
| Anti-RNA polymerate III | 1,170 | | 183 | |
| Positive | | 212 (18%) | | 33 (18%) |
| Negative | | 958 (82%) | | 150 (82%) |

**Supplementary Table S3.** Linear regression analysis of sociodemographic and disease characteristic associations with physical function (complete case analysis; N = 1,663)

| | **Physical Function Adjusted Regression Coefficient (95% CI)[a]** |
|---|---|
| Sociodemographic variables | |
| Age (years standardized) | **-0.74 (-1.16, -0.32)** |
| Male sex (reference = female) | **1.45 (0.24, 2.65)** |
| Years of education (years standardized) | **0.49 (0.09, 0.88)** |
| Single, divorced/separated, or widowed (reference = married or living as married) | -0.63 (-1.49, 0.22) |
| Non-White (reference = White) | -0.83 (-1.88, 0.22) |
| Country (reference = United States) | |
| Canada | 0.27 (-0.85, 1.38) |
| United Kingdom | -0.23 (-1.86, 1.41) |
| France | 0.69 (-0.34, 1.72) |
| Other (Australia, Mexico, Spain) | 1.68 (-0.11, 3.48) |
| Lifestyle variables and body mass index (BMI) | |
| Smoker (reference = non-smoker) | **-3.15 (-4.62, -1.67)** |
| Alcohol consumption (drinks per week standardized) | **0.53 (0.13, 0.93)** |
| BMI (standardized) | **-1.62 (-2.02, -1.21)** |
| Disease variables | |
| Years since first non-Raynaud's symptoms (years standardized) | -0.01 (-0.43, 0.42) |
| Diffuse subtype (reference = limited or sine) | **-1.13 (-2.06, -0.20)** |
| Gastrointestinal involvement (reference = no) | **-2.83 (-3.97, -1.69)** |
| Digital ulcers (reference = no) | **-1.52 (-2.71, -0.33)** |
| Tendon friction rubs (reference = never) | |
| Current | -0.30 (-1.54, 0.95) |
| Past | -0.21 (-1.61, 1.20) |
| Small joint contractures (reference = none or mild) | |
| Moderate | **-1.83 (-2.99, -0.66)** |
| Severe | **-2.14 (-3.89, -0.38)** |
| Large joint contractures (reference = none or mild) | |
| Moderate | **-2.45 (-4.01, -0.89)** |
| Severe | -2.01 (-4.68, 0.67) |
| History of SSc renal crisis (reference = no) | -0.45 (-2.54, 1.65) |
| Interstitial lung disease (reference = no) | **-1.61 (-2.47, -0.76)** |
| Pulmonary arterial hypertension (reference = no) | **-4.79 (-6.26, -3.32)** |
| Overlap syndromes | |
| Systemic lupus erythematosus (reference = no) | -1.19 (-3.61, 1.22) |
| Rheumatoid arthritis (reference = no) | **-2.46 (-4.43, -0.50)** |
| Sjogren's syndrome (reference = no) | -1.31 (-2.79, 0.17) |
| Autoimmune thyroid disease (reference = no) | -0.19 (-1.72, 1.34) |
| Idiopathic inflammatory myositis (reference = no) | **-2.52 (-4.30, -0.74)** |
| Primary biliary cirrhosis (reference = no) | 0.00 (-3.09, 3.09) |

Adjusted $R^2$ = 0.18

[a]All regression coefficients are unstandardized. Standardized variables calculated by subtracting raw scores from mean and dividing by standard deviation. Bolded results are statistically significant (P<0.05).

**Supplementary Table S4.** Linear regression analysis of sociodemographic and disease characteristic associations with physical function (sensitivity analysis; N = 2,385; adding pruritus and pain severity)

| | **Physical Function Adjusted Regression Coefficient (95% CI)[a]** |
|---|---|
| Sociodemographic variables | |
| Age (years standardized) | **-0.78 (-1.08, -0.49)** |
| Male sex (reference = female) | 0.59 (-0.25, 1.44) |
| Years of education (years standardized) | -0.14 (-0.42, 0.15) |
| Single, divorced/separated, or widowed (reference = married or living as married) | -0.33 (-0.93, 0.27) |
| Non-White (reference = White) | 0.00 (-0.75, 0.75) |
| Country (reference = United States) | |
| Canada | 0.38 (-0.38, 1.15) |
| United Kingdom | -0.08 (-1.10, 0.95) |
| France | **0.83 (0.09, 1.56)** |
| Other (Australia, Mexico, Spain) | 0.53 (-0.89, 1.94) |
| Lifestyle variables and body mass index (BMI) | |
| Smoker (reference = non-smoker) | **-1.35 (-2.42, -0.28)** |
| Alcohol consumption (drinks per week standardized) | 0.18 (-0.11, 0.46) |
| BMI (standardized) | **-0.79 (-1.07, -0.51)** |
| Disease variables | |
| Years since first non-Raynaud's symptoms (years standardized) | -0.02 (-0.33, 0.29) |
| Diffuse subtype (reference = limited or sine) | **-1.36 (-2.02, -0.69)** |
| Gastrointestinal involvement (reference = no) | **-1.55 (-2.35, -0.76)** |
| Digital ulcers (reference = no) | **-0.91 (-1.72, -0.09)** |
| Tendon friction rubs (reference = never) | |
| Current | -0.04 (-0.99, 0.92) |
| Past | 0.07 (-0.98, 1.12) |
| Small joint contractures (reference = none or mild) | |
| Moderate | -0.63 (-1.46, 0.20) |
| Severe | -1.17 (-2.40, 0.07) |
| Large joint contractures (reference = none or mild) | |
| Moderate | -1.02 (-2.13, 0.09) |
| Severe | -1.41 (-3.15, 0.33) |
| History of SSc renal crisis (reference = no) | -1.24 (-2.63, 0.15) |
| Interstitial lung disease (reference = no) | **-1.72 (-2.34, -1.10)** |
| Pulmonary arterial hypertension (reference = no) | **-3.69 (-4.71, -2.67)** |
| Pruritus (standardized) | **-0.68 (-1.00, -0.37)** |
| Pain severity (standardized) | **-4.55 (-4.87, -4.24)** |
| Overlap syndromes | |
| Systemic lupus erythematosus (reference = no) | **-1.81 (-3.53, -0.09)** |
| Rheumatoid arthritis (reference = no) | -0.50 (-1.77, 0.77) |
| Sjogren's syndrome (reference = no) | -0.21 (-1.31, 0.89) |
| Autoimmune thyroid disease (reference = no) | -0.11 (-1.27, 1.05) |
| Idiopathic inflammatory myositis (reference = no) | **-1.78 (-3.05, -0.50)** |
| Primary biliary cirrhosis (reference = no) | 0.64 (-1.40, 2.68) |

Adjusted $R^2$ = 0.43

[a]All regression coefficients are unstandardized. Standardized variables calculated by subtracting raw scores from mean and dividing by standard deviation. Bolded results are statistically significant (P<0.05).

**Supplementary Table S5.** Linear regression analysis of sociodemographic and disease characteristic associations with physical function (sensitivity analysis; N = 2,385; replacing disease subtype with continuous Modified Rodnan Skin Scores)

| | **Physical Function Adjusted Regression Coefficient (95% CI)[a]** |
|---|---|
| Sociodemographic variables | |
| Age (years standardized) | **-0.71 (-1.07, -0.35)** |
| Male sex (reference = female) | **1.29 (0.28, 2.31)** |
| Years of education (years standardized) | **0.36 (0.02, 0.71)** |
| Single, divorced/separated, or widowed (reference = married or living as married) | **-0.74 (-1.47, -0.01)** |
| Non-White (reference = White) | -0.76 (-1.66, 0.15) |
| Country (reference = United States) | |
| Canada | -0.14 (-1.07, 0.78) |
| United Kingdom | -1.19 (-2.44, 0.06) |
| France | 0.68 (-0.21, 1.56) |
| Other (Australia, Mexico, Spain) | **1.51 (-0.19, 3.21)** |
| Lifestyle variables and body mass index (BMI) | |
| Smoker (reference = non-smoker) | **-3.13 (-4.42, -1.85)** |
| Alcohol consumption (drinks per week standardized) | 0.80 (0.46, 1.15) |
| BMI (standardized) | **-1.40 (-1.73, -1.06)** |
| Disease variables | |
| Years since first non-Raynaud's symptoms (years standardized) | -0.16 (-0.53, 0.21) |
| Modified Rodnan Skin Score (standardized) | **-0.99 (-1.41, -0.58)** |
| Gastrointestinal involvement (reference = no) | **-2.59 (-3.55, -1.64)** |
| Digital ulcers (reference = no) | -1.78 (-2.77, -0.79) |
| Tendon friction rubs (reference = never) | |
| Current | -0.50 (-1.66, 0.65) |
| Past | -0.31 (-1.51, 0.89) |
| Small joint contractures (reference = none or mild) | |
| Moderate | **-1.78 (-2.78, -0.79)** |
| Severe | **-1.41 (-2.91, 0.09)** |
| Large joint contractures (reference = none or mild) | |
| Moderate | **-1.82 (-3.16, -0.48)** |
| Severe | -2.07 (-4.20, 0.06) |
| History of SSc renal crisis (reference = no) | -0.41 (-2.09, 1.28) |
| Interstitial lung disease (reference = no) | **-1.47 (-2.23, -0.72)** |
| Pulmonary arterial hypertension (reference = no) | **-3.65 (-4.84, -2.45)** |
| Overlap syndromes | |
| Systemic lupus erythematosus (reference = no) | -1.67 (-3.73, 0.38) |
| Rheumatoid arthritis (reference = no) | **-2.34 (-3.88, -0.80)** |
| Sjogren's syndrome (reference = no) | -0.98 (-2.33, 0.36) |
| Autoimmune thyroid disease (reference = no) | 0.07 (-1.33, 1.47) |
| Idiopathic inflammatory myositis (reference = no) | **-2.19 (-3.73, -0.66)** |
| Primary biliary cirrhosis (reference = no) | 0.96 (-1.48, 3.41) |

Adjusted $R^2 = 0.17$

[a]All regression coefficients are unstandardized. Standardized variables calculated by subtracting raw scores from mean and dividing by standard deviation. Bolded results are statistically significant (P<0.05).

**Supplementary Table S6.** Linear regression analysis of sociodemographic and disease characteristic associations with physical function (sensitivity analysis; N = 2,385; adding SSc-related antibodies)

| | **Physical Function Adjusted Regression Coefficient (95% CI)[a]** |
|---|---|
| Sociodemographic variables | |
| Age (years standardized) | **-0.69 (-1.06, -0.33)** |
| Male sex (reference = female) | **1.38 (0.37, 2.40)** |
| Years of education (years standardized) | **0.40 (0.06, 0.74)** |
| Single, divorced/separated, or widowed (reference = married or living as married) | **-0.73 (-1.46, -0.01)** |
| Non-White (reference = White) | -0.79 (-1.70, 0.12) |
| Country (reference = United States) | |
| Canada | -0.27 (-1.20, 0.66) |
| United Kingdom | -1.15 (-2.40, 0.09) |
| France | 0.52 (-0.39, 1.42) |
| Other (Australia, Mexico, Spain) | 1.40 (-0.31, 3.11) |
| Lifestyle variables and body mass index (BMI) | |
| Smoker (reference = non-smoker) | **-2.99 (-4.29, -1.70)** |
| Alcohol consumption (drinks per week standardized) | **0.81 (0.47, 1.16)** |
| BMI (standardized) | **-1.39 (-1.73, -1.05)** |
| Disease variables | |
| Years since first non-Raynaud's symptoms (years standardized) | -0.06 (-0.43, 0.31) |
| Diffuse subtype (reference = limited or sine) | **-1.33 (-2.23, -0.43)** |
| Gastrointestinal involvement (reference = no) | **-2.58 (-3.54, -1.62)** |
| Digital ulcers (reference = no) | **-2.08 (-3.07, -1.09)** |
| Tendon friction rubs (reference = never) | |
| Current | -0.82 (-1.97, 0.34) |
| Past | -0.12 (-1.35, 1.11) |
| Small joint contractures (reference = none or mild) | |
| Moderate | **-1.92 (-2.92, -0.91)** |
| Severe | **-1.72 (-3.22, -0.23)** |
| Large joint contractures (reference = none or mild) | |
| Moderate | **-2.04 (-3.38, -0.70)** |
| Severe | **-2.67 (-4.80, -0.53)** |
| History of SSc renal crisis (reference = no) | -0.09 (-1.81, 1.64) |
| Interstitial lung disease (reference = no) | **-1.78 (-2.59, -0.96)** |
| Pulmonary arterial hypertension (reference = no) | **-3.72 (-4.92, -2.52)** |
| Overlap syndromes | |
| Systemic lupus erythematosus (reference = no) | -1.63 (-3.69, 0.43) |
| Rheumatoid arthritis (reference = no) | **-2.07 (-3.63, -0.52)** |
| Sjogren's syndrome (reference = no) | -0.88 (-2.21, 0.44) |
| Autoimmune thyroid disease (reference = no) | 0.00 (-1.41, 1.40) |
| Idiopathic inflammatory myositis (reference = no) | **-1.97 (-3.53, -0.42)** |
| Primary biliary cirrhosis (reference = no) | 0.85 (-1.61, 3.31) |
| SSc-related antibodies | |
| Antinuclear antibodies (reference = negative) | 0.59 (-1.00, 2.17) |
| Anti-centromere (reference = negative) | 0.17 (-0.80, 1.15) |
| Anti-topoisomerase I [Scl70] (reference = negative) | **0.96 (0.04, 1.89)** |
| Anti-RNA polymerate III (reference = negative) | -0.51 (-1.69, 0.66) |

Adjusted $R^2$ = 0.17

[a]All regression coefficients are unstandardized. Standardized variables calculated by subtracting raw scores from mean and dividing by standard deviation. Bolded results are statistically significant ($P<0.05$).
